# A clinical microscopy dataset to develop a deep learning diagnostic test for urinary tract infection

**DOI:** 10.1101/2023.09.19.23295802

**Authors:** Natasha Liou, Trina De, Adrian Urbanski, Catherine Chieng, Qingyang Kong, Anna L David, Rajvinder Khasriya, Artur Yakimovich, Harry Horsley

## Abstract

Urinary tract infection (UTI) is a common disorder. Its diagnosis can be made by microscopic examination of voided urine for cellular markers of infection. This manual technique is technically difficult, time-consuming and prone to inter-observer errors. The application of computer vision to this domain has been slow due to the lack of a clinical image dataset from UTI patients. We present an open dataset containing 300 images and 3,562 manually annotated urinary cells labelled into seven classes of clinically significant cell types. It is an enriched dataset acquired from the unstained and untreated urine of patients with symptomatic UTI using a simple imaging system. We demonstrate that this dataset can be used to train a Patch U-Net, a novel deep learning architecture with a random patch generator to recognise urinary cells. Our hope is that with this dataset UTI diagnosis will be made possible in nearly all clinical settings by using a simple imaging system which leverages advanced machine learning techniques.

## Background & Summary

UTI can often be clinically identified by the presence of lower urinary tract symptoms (LUTS), with the classical symptoms being burning or pain on urination and frequency of urination. UTIs are the most common bacterial infection in humans with the potential to become a recurrent / chronic infection and lead to life-threatening sepsis^1^. Women are not only at increased risk of UTI, but also more likely to develop complicated infections^2^. Not surprisingly, UTIs are associated with a substantial health and economic burden^3^ and the prevalence of antibiotic prescriptions and hospital admissions related to urine infections is on the rise^4,5^.

Rapid identification of infection and timely administration of antimicrobial treatment can prevent adverse complications. Point-of-care testing (POCT), tests which are performed at the bedside at the time and place of patient care, is the preferred diagnostic practice^6^. However, the current tests routinely available are inadequate to detect UTI. Urine dipstick is one such POCT, but fails to detect complicated and recurrent infection^7^. Midstream urine culture is the supposed gold standard, but cannot be done at the bedside, underreports pathogenic microbes, and does not describe the host response to infection nor correctly report the urobiome^8^. Without an accurate POCT, clinicians are ill equipped to diagnose infections, thus contributing to inappropriate antibiotic use and potentially driving antimicrobial resistance^9^.

Urine microscopy and identification of urinary cells from freshly voided urine is an alternative POCT with greater sensitivity than both aforementioned methods. The presence of white blood cells (WBC, or pyuria) in an unspun, unstained specimen of urine is particularly predictive of a UTI^10^. Here, WBC quantification must be performed at the bedside, soon after the void, to prevent cell degradation and subsequent underestimation of pyuria^11^. The presence of epithelial cells (EPC) is also suggestive of infection as urinary epithelial cells are actively involved in antibacterial activities^12^. EPC parasitised by bacteria, in humans and mice, have been shown to be jettisoned from the urothelium in an effort to lower bacterial load^13,14^.

Urinary microscopy measures and explores the host immune response and, therefore, reflects the underlying pathophysiological state of the host’s urinary tract. Use of this test has been shown to improve patient outcomes^15^. Pain, storage, and voiding symptoms have been found to be the most reliable predictors of microscopic pyuria, and in turn correlate with measures of quality of life. In our experience, treating patients with chronic LUTS, we find that peak symptoms coincide with peak cell counts (Figure 1). Unfortunately, while the benefits of urine microscopy have clear clinical benefits, it requires the time and manual labour of an experienced microscopist thus limiting its availability as a POCT to highly specialised clinics in well-developed countries^11^.

**Figure 1:**
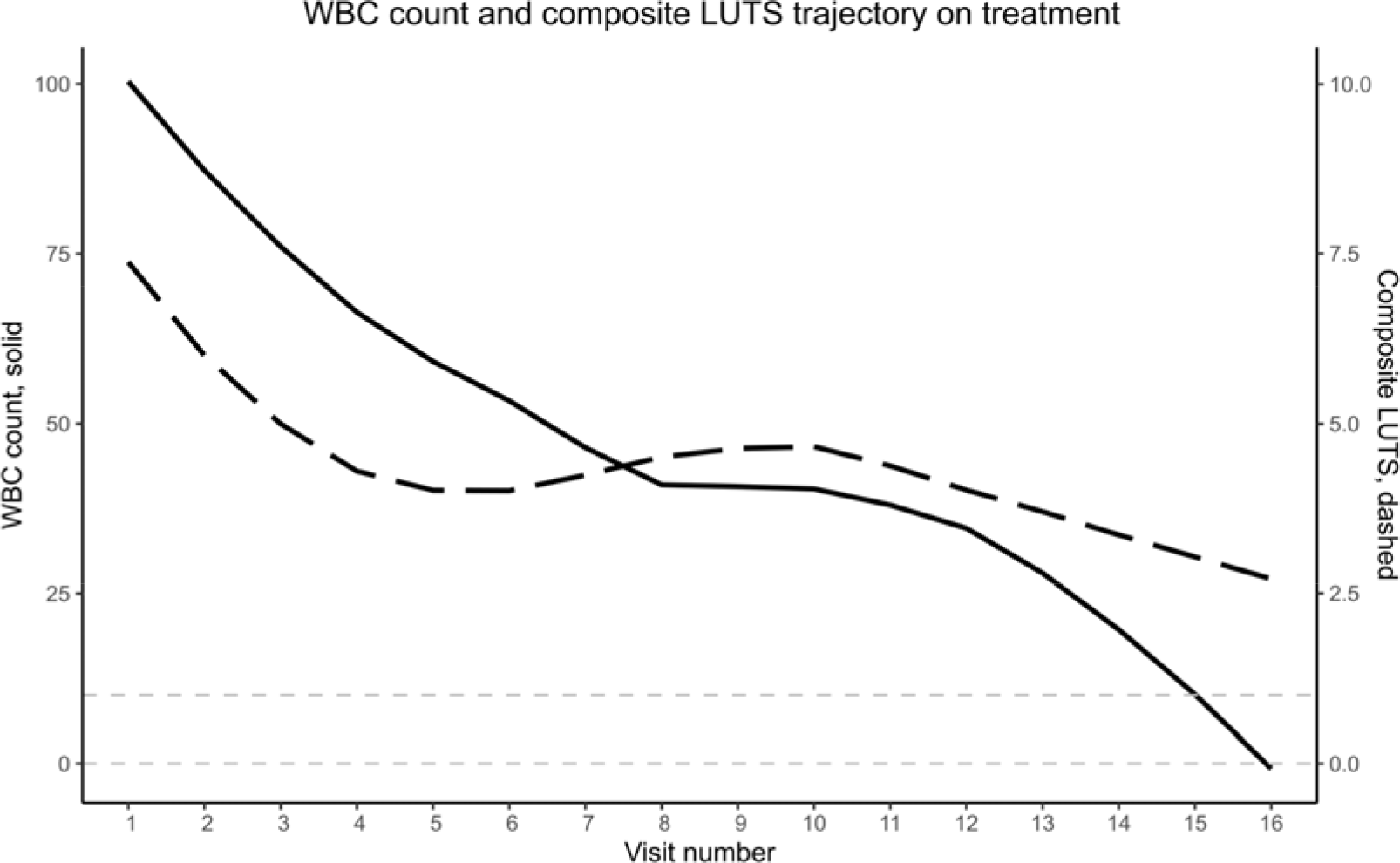
WBC count and composite LUTS trajectory on treatment. WBC count (solid line) as measured by urine microscopy correlates with a composite score of LUTS (dashed line) over the course of antimicrobial treatment. By the final clinical attendances, WBC count drops within a physiological WBC range as marked by the pale dashed grey lines.

Machine learning in biomedical imaging is increasingly used as an adjunct to enhance or automate conventional diagnostics. However, a clinically relevant and representative dataset of urinary cells indicative of infection does not currently exist. This is due to the wide range and often equivocal nature of cellular content in symptomatic patients. While these difficulties might be overcome by the application of histological stains^16^, such a model could no longer be offered as a POCT as the workflow would require access to specialised equipment found in large, centralised laboratories away from the point of patient care. Moreover, such advanced equipment and techniques are likely only to be available in well-developed countries. There is no open dataset of high-quality urinary cells annotated for the analytical task of UTI detection to date.

We have produced an open image dataset of urinary cells which can be used to identify markers of infection using machine learning techniques. Our image dataset of voided urine is clinically representative of patients with known urine infection. Unlike other cellular image datasets, cell identification techniques such as histological staining have not been deployed and therefore minimal laboratory processing is required. This was purposefully done with the ambition of creating an accurate POCT using a simple imaging system which leverages machine learning.

## Methods

### Clinical samples

300 urine samples were randomly obtained from patients with symptomatic UTI from the Whittington Health NHS Trust in London, UK. LUTS data was collected using a validated 39-question inventory grouped into pain, urgency, voiding, and stress symptoms and assessed in binary yes or no response (Supplementary File 1). Frequency of urination and incontinence during the day and night was also assessed.

### Data acquisition

Urine samples were collected as natural voids and processed on-site within one hour to limit cellular degradation. Brightfield microscopic examination (Olympus BX41F microscope frame, U-5RE quintuple nosepiece, U-LS30 LED illuminator, U-AC Abbe condenser) was performed using a x20 objective (Olympus PLCN20x Plan C N Achromat 20x/0.4). A disposable haemocytometer (C Chip™) was used for enumeration of red cells (RBC), white cells (WBC), epithelial cells (EPC), and the presence of other cellular content per 1 μl of urine by two experienced microscopists.

Images were acquired using the aforementioned brightfield microscope coupled to a digital scientific colour camera (Infinity 3S-1UR, Teledyne Lumenera) via a 0.5X C-mount. Images were taken in 16-bit colour in 1392 × 1040 .tif format using Micromanager software^17^. Daily Kohler illumination and global white balance was performed to ensure consistency in image acquisition. An enriched dataset approach was taken to maximise urinary cellular content in the acquired images. Such data curation was also necessary to attenuate object sparsity.

### Dataset annotation

300 images were acquired and manually annotated by first identifying cells of interest as a binary semantic segmentation task. Individual pixels were dichotomously labelled as either informative objects, foreground, or non-informative background. Non-informative background was further constrained by including unidentifiable cells, such as debris or grossly out of focus particles. Binary annotation was initially performed using ilastik^18^, an open source software using a random forest classifier for pixel classification, then manually refined at the pixel level to ensure accurate segmentation. This produced a binary mask in 1392 × 1040 .tif format with values [0,1] for each corresponding raw colour image.

Cells of interest were subsequently labelled manually by two expert microscopists into one of seven clinically significant multi-class categories: rods, RBC/WBC, yeast, miscellaneous, single EPC, small EPC sheets, and large EPC sheets. This produced a multi-class mask in 1392 × 1040 .tif format with integral values between [0,7].

### Data preprocessing

First, the image was rescaled according to the scale factor either 0.2, 0.3, 0.5 or 1, and thereby, if applicable, decreasing its resolution. This allowed the model to analyse a larger area while keeping the patch size uniform, an valuable strategy in the case of sparse data. Next, 256×256 patches were cut from a random region in the image. Finally, all values within the patch were rescaled to fall within the range of [-1,1] by performing the following operations: divide by 255, the highest potential value, then multiply by 2, and finally subtract 1. In the case of training data, random vertical and horizontal flips were performed to increase the variation in the data and encourage model generalisation.

### Patch U-Net architecture

Generally, we followed the architectures described here^19,20^. There were, however, a few notable changes. Firstly, we added instance normalisation layers. Secondly, we made the size of the network scalable by specifying the number of channels produced by the initial convolutional layer.

Similarly to the architecture proposed by Ronneberger and colleagues^19^, our network consisted of an encoder (contracting) and decoder (expansive) path. A critical component of the network was the convolutional block, which consisted of repeated applications of 3×3 convolutions, each followed by batch normalisation and rectified linear unit (ReLU). The contracting path consisted of 5 convolutional blocks, each followed by an instance normalisation layer and a 2×2 max pooling operation with stride 2 for downsampling. After each downsampling step, we doubled the number of feature channels.

Every step in the expansive path consisted of an upsampling of the feature map followed by a 2×2 convolution which halved the number of feature channels, followed by a convolutional block and instance normalisation layer.

### Loss functions

The loss function was computed by a pixel-wise sigmoid over the final feature map with the combined binary cross entropy and Dice coefficient loss function (see Equation (5)).

The sigmoid function is defined as:

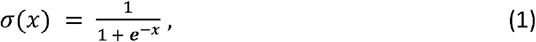

and casts the prediction values into (0,1) range. Let’s define *Y* as ground truth, *Ŷ* as model prediction, and *N* as the number of pixels. The cross-entropy penalises the deviation from the ground truth at each position using:

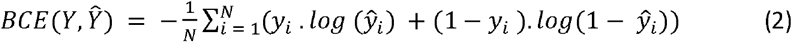

This is combined with dice loss, which is defined as:

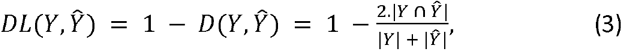

where *D* (*Y, Ŷ*) is defined in Equation (5). The final loss function is defined as:

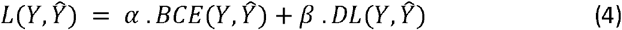

It is calculated across the batch to make it more stable. In our experiments, we set *α* = *β* = 1.

### Batch generation

To prepare batches of training data for the Patch U-Net, full-scale images were dynamically pre-processed into patches of 256×256 pixels during the training. Training of the Patch U-Net was performed on mini-batches of such patches. Given the sparsity of the objects in the images, a procedure evaluating emptiness of the image was devised. As a result, for each mini-batch, patches were generated using the following procedure:

1. Choose a random value as

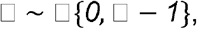

where □ is the height of the images
2. Choose a random value as

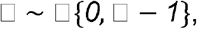

where *W* is the width of the images and *U* represents a Discrete Uniform Distribution^21^
3. Get patches from the image *X* as follows:

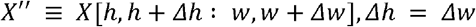
4. Get batches of patches as above as follows:

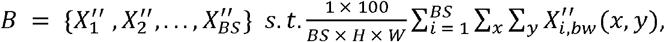

where *BS* is the batch size and 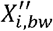 is the binary mask corresponding to the *i*^*th*^ image patch in the batch *B*.

### Metrics

To evaluate model performance during training we employed the Sørensen–Dice coefficient^22,23^ which measures the ratio between the area of overlap and the total number of pixels classified as foreground in both images and is described by Equation (1) :

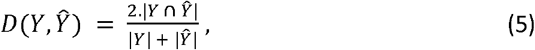

where *Ŷ* is the segmentation mask returned by the model, and *Y* is the ground truth.

### Training evaluation

During training, the performance on both train and validation sets was calculated mini-batch wise where *b, h, w* respectively correspond to the index of the sample in a mini-batch and the position of a pixel in the sample respectively.

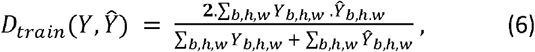

Such an approach allowed us to address the sparse patches, i.e. patches where only a few pixels were marked as foreground. Such patches could contribute unrealistically high performance, should the metric be calculated in a sample-wise manner. In contrast, our approach allowed us to alleviate such circumstances, ensuring better training performance.

### Testing evaluation

For the final evaluation, we opted to emulate the real-world inference, and thus the metrics were computed image-wise. Since our model was patch-based, each image was split into patches prior to inputting into the model. To avoid potential issues at the edges of each patch, inference was performed on overlapping patches. Next, predictions were combined into a final mask by means of taking maximum from overlapping regions.

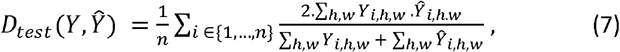

where *h, w* have the same definition as in Equation (6) and *i* is the image index in the test set.

### Model implementation

#### Optimiser

To optimise the model’s parameters, we employed the Adam optimiser^24^ with an initial learning rate of 0.001. Then, we decreased the learning rate according to an exponential schedule with a decay rate of 0.95 every 50 epochs.

### Regularisation

To prevent overfitting, we employed the following regularisation technique: L2 weight decay. L2 weight decay with a coefficient of 0.0001 was used to penalise large weights and encourage the model to arrive at sparse solutions.

### Training procedure

Our training process consisted of 750 epochs for each experiment giving the model sufficient time to converge, each epoch containing 1000 random samples. We used a batch size of 50, and the model’s parameters were updated with mini-batch gradient descent.

### Hardware and software setup

The model was built in Python 3.10.8. TensorFlow, a library developed to solve deep learning problems, was incorporated to increase model scalability, speed, and accuracy. Keras was used as a Python interface to TensorFlow. The following libraries and their required versions used in our network were as follows: keras 2.6.0, keras-preprocessing 1.1.2, numpy 1.19.5, tensorboard 2.6.0, tensorflow 2.6.0, scikit-image 0.18.1, tqdm, scipy, seaborn, and scikit-learn. Experiments were conducted on the following machines: MacBook Pro Apple with M1 Max Chip with 10-core CPU and 32-core GPU, HPC Hemera at HZDR on a Nvidia Tesla A100 GPU 40GB, HPC at ZIH TU Dresden on a NVIDIA A100-SXM4 Tensor Core-GPU.

### Ethics

Written informed consent was obtained from all participants in accordance with Good Clinical Practice guidance. Ethics was approved by Health Research Authority (HRA) and Health and Care Research Wales (HCRW) under “A prospective observational cohort study of the pathophysiology of urinary tract infection”, IRAS 295252, protocol number 143470, and REC reference 22/WA/0069.

## Data Records

### Demographics and symptomatology

Image dataset was obtained from urine samples of patients with symptomatic UTI. 300 patients (mean 42 ± 15 years, 95.1% female) were recruited over the study time period. Patients reported a total of 3 LUTS (IQR 1-6 symptoms), with pain being the predominant symptom (median pain score 2, IQR 0-3), followed by storage, voiding, and stress (Figure 2). Most samples contained WBC (median 6, IQR 2-22 WBC per 1 μl urine) and EPC (median 14, IQR 4-42 EPC per 1 μl urine), and were negative for RBC.

**Figure 2:**
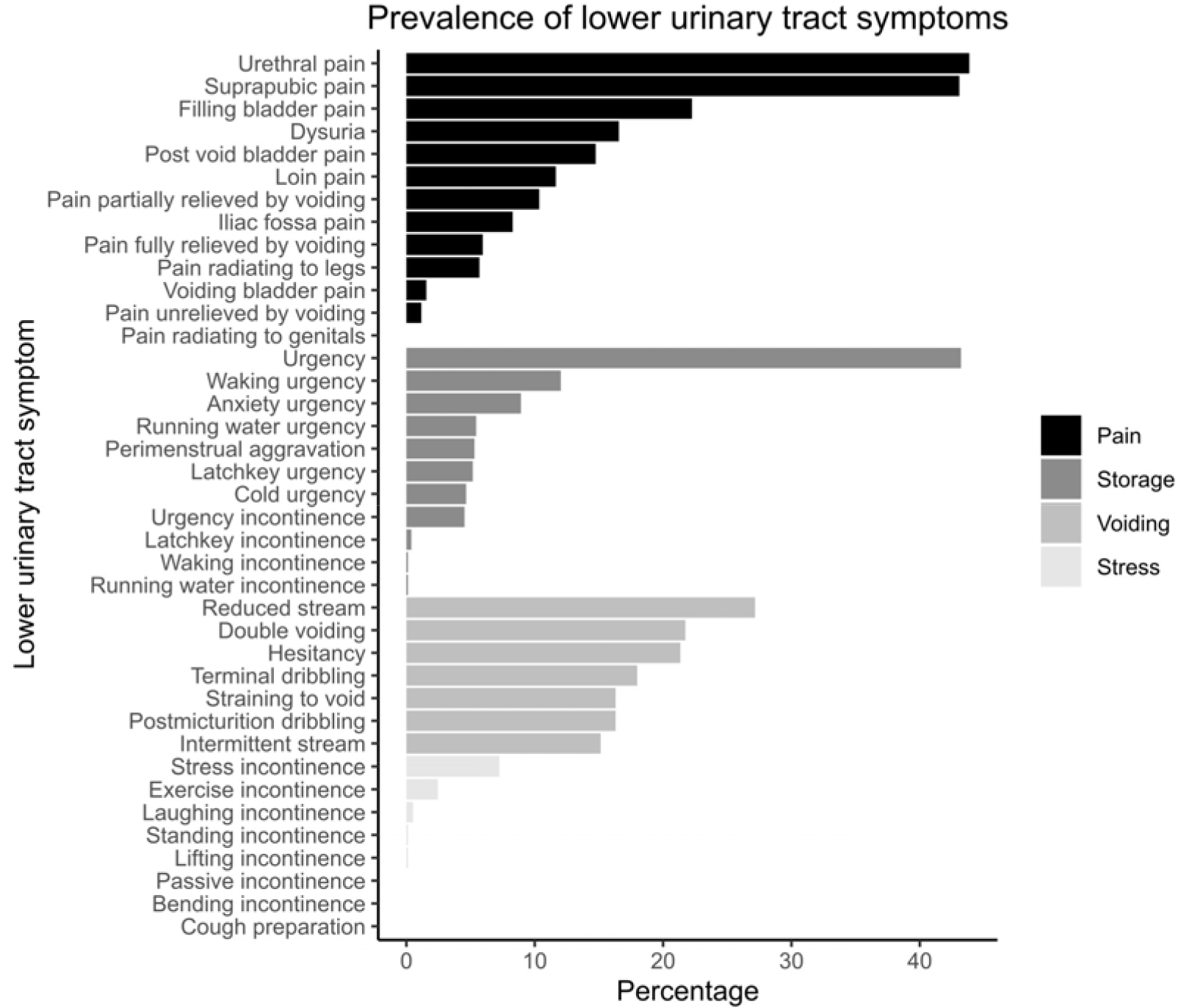
Prevalence of lower urinary tract symptoms (LUTS). Pain was the most common symptom associated with this cohort of patients, followed by storage and voiding. This is in keeping with the symptoms most predictive of microscopic pyuria and, in turn, UTI^15^.

### Binary and multi-class labels

300 images were manually annotated to produce 3,562 objects. All 3,562 objects were labelled into one of seven clinically distinct multi-class cell types: rods, RBC/WBC, yeast, miscellaneous, single EPC, small EPC sheets, and large EPC sheets (Table 1).

**Table 1:** Data structure.

| Folder | Files | Objects | Count | Pixel Values |
| --- | --- | --- | --- | --- |
| img | 300 | Raw data |  | 0-65535 |
| bin_mask | 300 | Background/Foreground |  | 0/1 |
| mult_mask | 300 | Background/Class |  | 0 |
|  |  | Rod | 1697 | 1 |
|  |  | RBC/WBC | 1056 | 2 |
|  |  | Yeast | 41 | 3 |
|  |  | Miscellaneous | 550 | 4 |
|  |  | Single EPC | 182 | 5 |
|  |  | Small EPC sheet | 26 | 6 |
|  |  | Large EPC sheet | 10 | 7 |
|  |  | Total | 3562 |  |

These classes were chosen due to their clinical significance. Coliform bacteria are frequently implicated in UTI pathogenesis and are rod-shaped with each cell unit measuring 0.25 - 1.0 μm in width and 2.0 μm in length. These bacteria can elongate up to 15μm to produce a filamentous morphology, a phenomenon often associated with bacterial pathogenicity in the urinary tract^25,26^. Yeast (most commonly of the *Candida* species) are also seen in urine, and may represent a commensal organism or infectious pathogen^27^. Their size is dependent on their mitotic state, and in certain states, may be confused with erythrocytes. RBC and WBC, haematuria and pyuria respectively, are cellular indicators of infection^28,29^. EPC are often seen as individual cells or sheets of cells. A powerful mechanism to rapidly reduce bacterial load is to shed the superficial bladder epithelium invaded by bacteria^12^. The presence of large EPC sheets may therefore indicate more widespread infection hence more extreme cellular exfoliation. Work is ongoing to further subtype the aforementioned classes (e.g. distinct WBC populations such as neutrophils, macrophages and lymphocytes) and annotate new classes (e.g. cocci, another bacterial morphology).

### Data structure

The dataset is organised into three root folders: image, binary mask, and multi-class mask (Table 1). Each folder has 300 files in .tif format and labelled incrementally.

### Data storage

The dataset is publicly available at the Rodare data repository^30^. Images were captured at the clinic and anonymised using an allocated study number. Images were stored in secure UCL storage. All patient data and manual microscopy reports were entered on an encrypted database on a secure server in compliance with General Data Protection Regulation. This clinical database is NHS approved and procured, and regularly backed up.

## Technical Validation

### Binary semantic segmentation using a neural network with random patch generator

To evaluate the applicability of the dataset to deep-learning-based image segmentation, we developed a patch-based U-Net (Patch U-Net) similar to several other architectures proposed previously^20,31^ to perform urinary cell identification by binary semantic segmentation. The architecture of the proposed model incorporates a unique random patch generator (Figure 3a) to produce multiple input and output patches at different resolutions in the requisite square-shaped U-net dimensions for data augmentation. The image and binary mask components of the dataset were equally and randomly split into train, validate, and test subsets with 100 images each. We chose this data split, as opposed to the conventional 70/20/10 split, to mitigate potential underrepresentation of certain cell types that are morphologically distinct, sparse, and yet significant.

**Figure 3:**
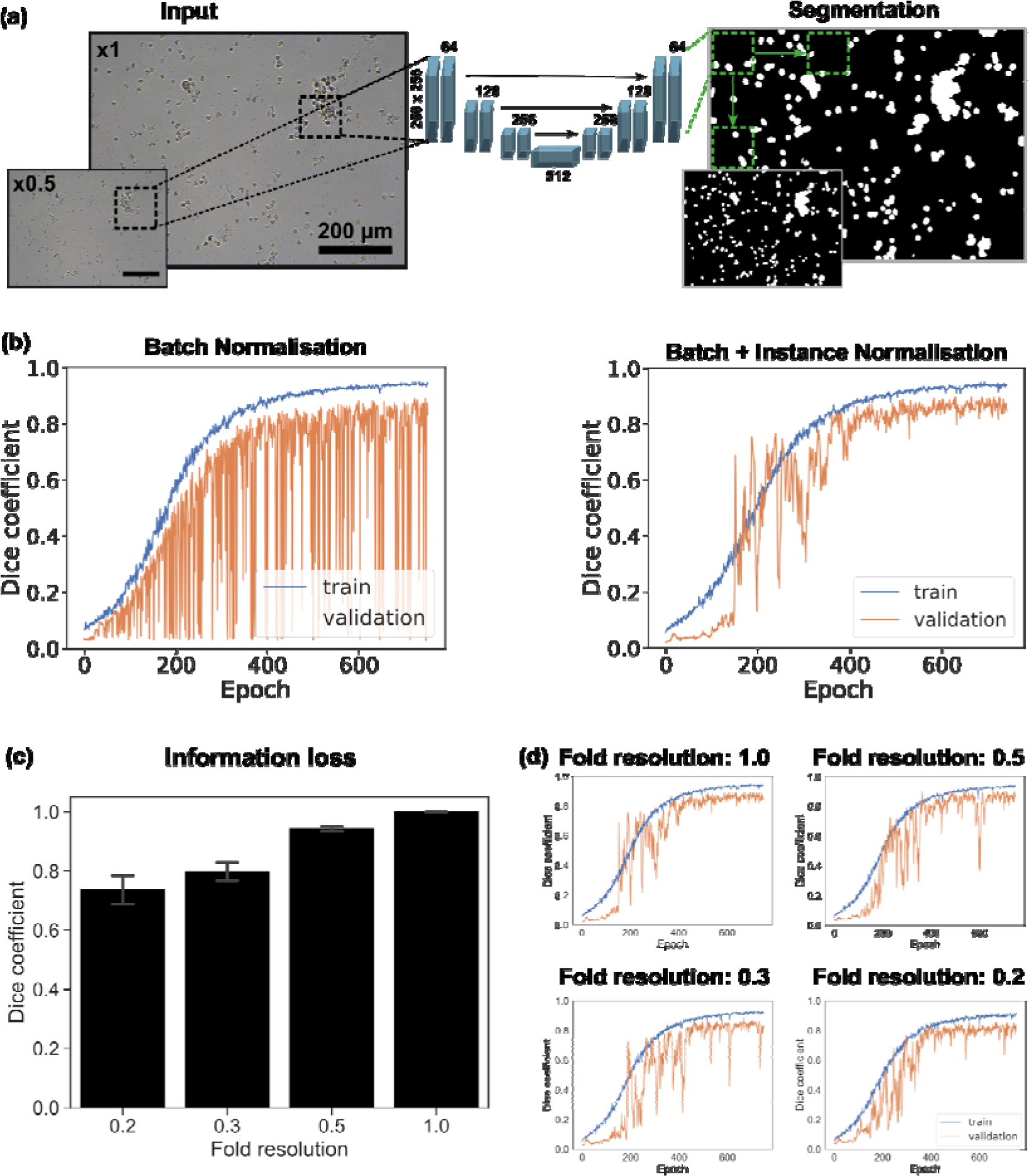
Binary Segmentation using Patch U-Net. (a) Patch U-Net architecture with patches as input and output resolutions 1.0 and 0.5, (b) Training performance with batch normalisation only (left) and combined batch and instance normalisation (right), (c) Information loss measured as Dice coefficient (± standard deviation) between original and downscaled-upscaled images, (d) Training and validation performance using original images at full resolution 1.0 and reduced resolutions at scale factors of 0.5, 0.3 and 0.2.

Patch U-Net, which processes patches rather than whole images, was employed since preserving resolution was critical for detecting small objects such as bacteria. However, the dimensions of our input images were (1392, 1040, 1) making it computationally expensive to process entire images. Employing a Patch U-Net was also effective since our dataset is sparse in nature. Thus, the model can converge faster when shown data that is relevant for semantic segmentation rather than the background. For this a filter was applied to the generated patches, where a batch of patches of shape (batch size, 256, 256, 1) was used for training only when a specific criteria (see section Batch generation - Methods) was satisfied.

### Impact of data normalisation on binary segmentation

During the initial stages of our experiments, an issue appeared involving the instability of validation accuracy during training. Periodically, the model displayed abnormal behaviour, classifying entire images as either foreground or background, resulting in a significant drop in accuracy. Although this behaviour tended to persist for only a few epochs, it raised concerns. To tackle this problem, instance normalisation layers were incorporated into our network architecture. These layers played a crucial role in preventing instance-specific mean and covariance shifts, thereby simplifying the learning process. This technique, introduced by Ulyanov et al.^32^, effectively alleviated the instability observed during training (Figure 3b).

Images used at scale factors 0.2, 0.3 and 0.5 were found to have unstable validation curves. Given the wide range of cellular content, batch normalisation alone was tested against an alternative method of feature scaling using combined batch and instance normalisation. By normalising on the level of individual images, variance was contained within each instance thus reducing network noise to produce more stable curves (Figure 3d).

### Impact of image resolution on binary segmentation

To increase context within the same patch size while maintaining the same computational complexity, we tested the effects of reduced image resolutions. Such an approach is widely used in computer vision to increase computational efficiency. Specifically, we considered the resolutions at scale factors of 0.2, 0.3, 0.5 and 1 of the original resolution, referred to as fold resolution in Figure 3. Notably, pixel information was lost during downscaling to and consequent upscaling from any scale factor other than 1 (Figure 3c). This loss of information was measured as the average Dice coefficient between original images at full resolution i.e. scale factor 1 and the corresponding images scaled down to a lower resolution. For example, images reduced to a factor of 0.2, 0.3 or 0.5 in scale were then scaled back up to full resolution. Pixel information was increasingly lost as the scale factor decreased as seen in Figure 3c. The impact of resolution should therefore be carefully considered for this dataset.

To investigate if training on low resolution images and conducting inference on high resolution images could serve as a viable alternative, we trained binary segmentation models on scale factors of 0.2, 0.3, 0.5 and 1. Figure 3d shows the training performance of our model using different images at scale factors of the original resolution to generate patches. Validation was performed on similar patches of the respective downscaling factor from the validation set. Once trained, inference was performed on the full resolution images from the test set (Table 2). Remarkably, results suggested that a model trained on images downsampled as high as factor 0.3 of the full resolution may be as effective in inference on full resolution images, as the model trained on full resolution images.

**Table 2:**
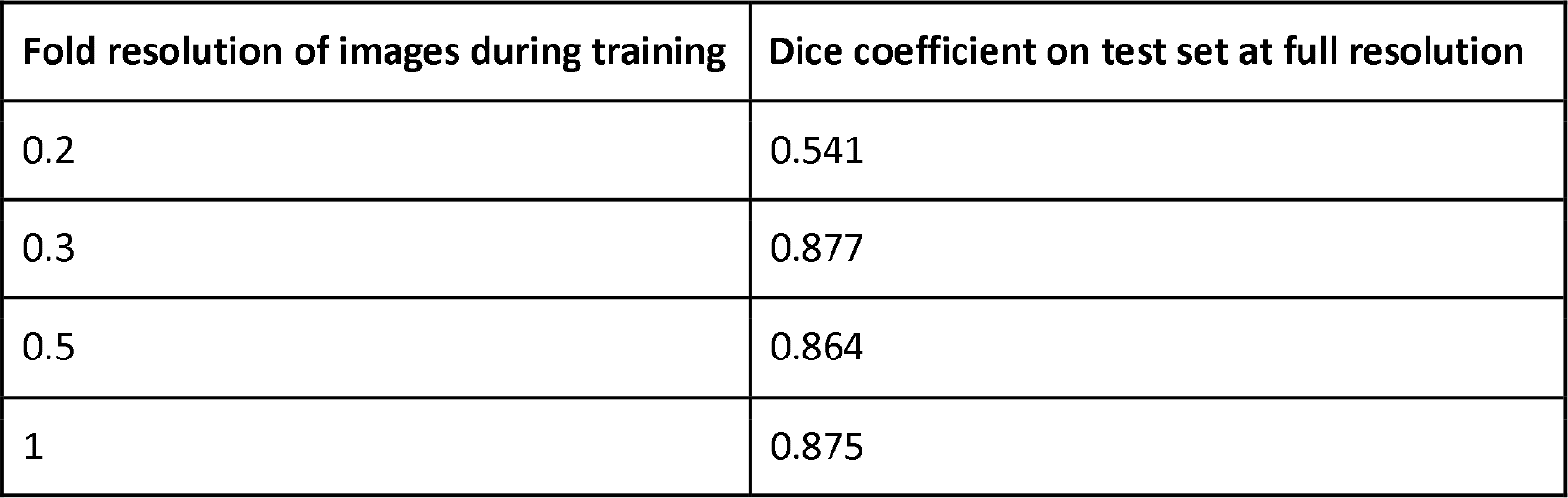
Patch U-Net performance on binary segmentation. Model training was performed at different image resolutions with model performance reported as a Dice coefficient on the test set at full image resolution.

### Multi-class morphological feature projection

To make our dataset applicable for computer vision tasks such as multi-class segmentation, object-detection and clustering, we annotated the binary masks into seven classes (see section Binary and multi-class labels - Data Records). Multi-class segmentation annotations can be translated into object-detection annotations. This can be achieved by treating binary masks as a set of connected components on a black background and obtaining bounding boxes of each connected component.

To examine properties of the multi-class objects in an interpretable manner, we evaluated projections of some morphological features which we found to be particularly distinct. Specifically we evaluated area (μm) and circularity (value between 0.0 to 1.0, where 1.0 represents a perfect circle). We also scaled these values further using a standard scaler^33,34^. For this we first obtained connected components from the pixel-level multi-class masks present in the dataset. Next, the connected components were projected as manually labelled classes using a scatterplot with both features scaled, and area additionally log transformed (Figure 4). Examples of each cell category are demonstrated in the legend.

**Figure 4:**
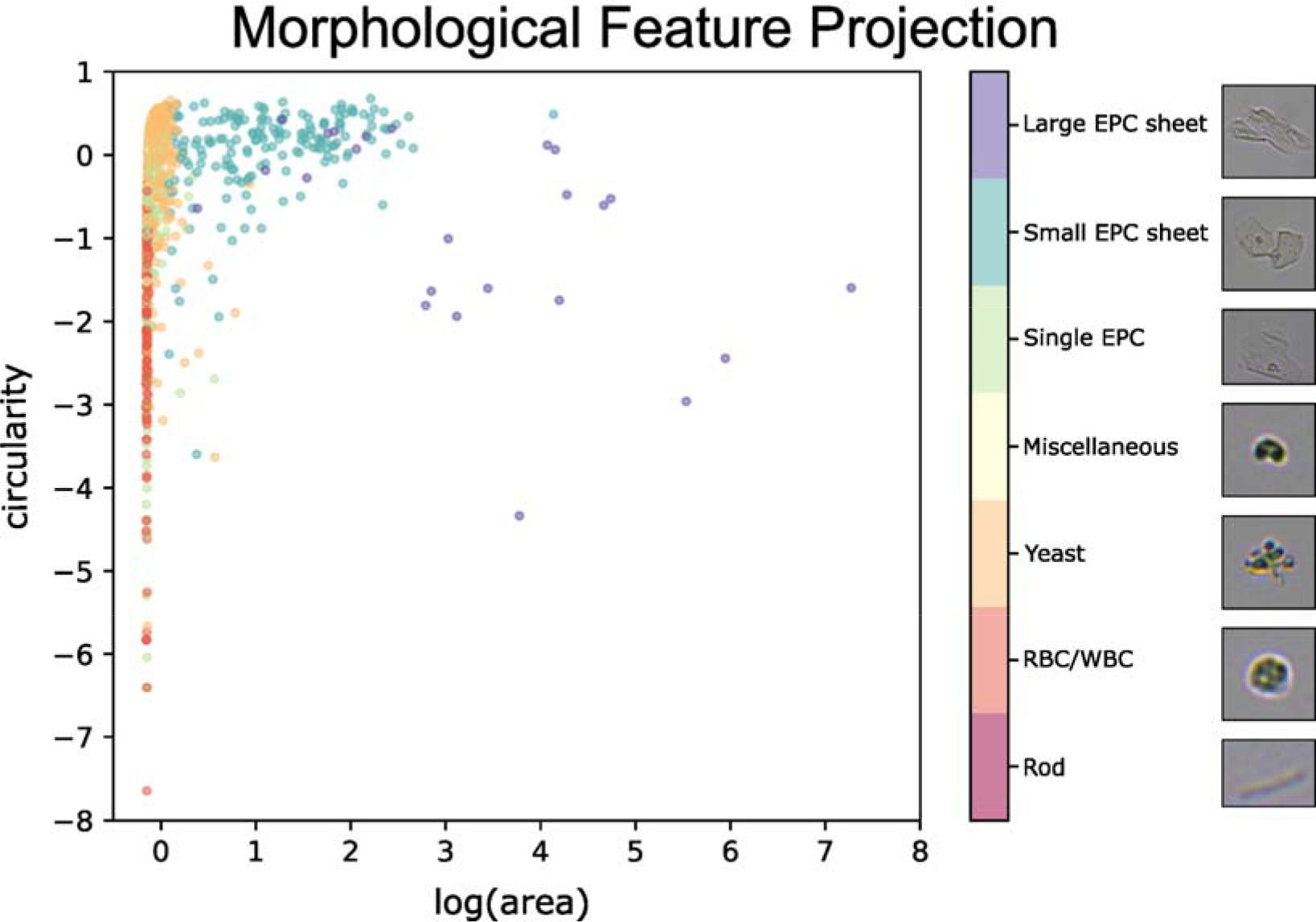
Morphological Feature Projection. 2D scatter plot of logarithm area vs. circularity measurements of connected components of interest extracted from binary segmentation (see Figure 3). Data points are coloured by class according to their corresponding cell type.

In summary, UTI is a rising global problem and current diagnostic tests perform poorly. Here, we present an annotated, clinically-relevant, image dataset to perform binary and multiclass segmentation and object detection. We demonstrate the applicability and real world potential of deep learning to this clinical problem by training a simple semantic segmentation model. Moreover, we explore and present the effect of data normalisation and image resolution on model performance. This proof-of-concept dataset represents the initial steps towards a more fit for purpose and equitable diagnostic test for UTI.

## Data Availability

All data produced in the present study are available upon reasonable request to the authors

http://doi.org/10.14278/rodare.2473

https://github.com/casus/UMOD

## Code Availability

All code is available from https://github.com/casus/UMOD under MIT open source licence.

## Acknowledgements

This work was partially funded by the Center for Advanced Systems Understanding (CASUS) which is financed by Germany’s Federal Ministry of Education and Research (BMBF) and by the Saxon Ministry for Science, Culture, and Tourism (SMWK) with tax funds on the basis of the budget approved by the Saxon State Parliament. It was also partially funded by the International Urogynecological Association (IUGA) through their Basic Science Research Grant and a philanthropic donor to the Bladder Infection and Immunity Group (BIIG).

## Author contributions

NL, TD, AU, CC, QK, AD, RK, AY and HH conceptualised the project. NL and HH performed sample preparations, image acquisition and data annotation. TD, AU, AY and NL wrote the program code. TD and AU designed the neural network and performed the computational experiments. NL, TD, AU, CC, QK, AD, RK, AY and HH wrote the manuscript.

