## Supplementary File 1 for "A clinical microscopy dataset to develop a deep learning diagnostic test for urinary tract infection"

Study Number: Date:

| Lower urinary tract symptoms (LUTS)<br>There are **39 questions**. Please circle your answer (Yes / No) | | |
|---|---|---|
| **Storage symptoms** | | **Circle** |
| 1. Urgency | Do you have to hurry to pass urine because you might wet yourself? | Y / N |
| 2. Urge incontinence | Do you hurry to pass urine but not make it in time? | Y / N |
| 3. Latchkey urgency | Do you find that you have to rush to pass urine when you put a key in the front door? | Y / N |
| 4. Latchkey urgency incontinence | If you have to rush to pass urine when you put a key in the front door, do you ever leak with this? | Y / N |
| 5. Waking urgency | Do you have to hurry to pass urine because you might wet yourself when you wake up in the morning? | Y / N |
| 6. Waking urge incontinence | If you have to hurry to pass urine when you wake up in the morning, do you ever wet yourself? | Y / N |
| 7. Running water urgency | Do you have to hurry to pass urine because you might wet yourself on hearing running water? | Y / N |
| 8. Running water urge incontinence | If you have to hurry to pass urine on hearing running water, do you ever wet yourself? | Y / N |
| 9. Cold urgency | Do you have to hurry to pass urine because you might wet yourself on exposure to cold? | Y / N |
| 10. Anxiety urgency | Do you have to hurry to pass urine because you might wet yourself on when you are worried or anxious? | Y / N |
| 11. Premenstrual aggravation | Do you have to hurry to pass urine because you might wet yourself, more around the time of your period? | Y / N |
| **Stress symptoms** | | **Circle** |
| 12. Cough sneeze incontinence | Do you leak on coughing or sneezing? | Y / N |
| 13. Exercise incontinence | Do you leak on exercise? | Y / N |
| 14. Laughing incontinence | Do you leak on laughing? | Y / N |
| 15. Passive incontinence | Do you leak for no good reason? | Y / N |
| 16. Bending incontinence | Do you leak on bending? | Y / N |
| 17. Standing incontinence | Do you leak on standing from sitting? | Y / N |
| 18. Lifting incontinence | Do you leak on lifting anything? | Y / N |
| 19. Pre-cough preparation | Do you get ready to avoid leaking when you are about to cough? | Y / N |

| Voiding symptoms | | Circle |
|---|---|---|
| 20. Hesitancy | Is to slow to start passing urine? | Y / N |
| 21. Reduced stream | Is the urinary stream reduced? | Y / N |
| 22. Intermittent stream | Does the stream stop and start? | Y / N |
| 23. Straining to void | Do you have to strain to pass urine? | Y / N |
| 24. Terminal dribbling | Does the stream dribble at the end? | Y / N |
| 25. Postvoid dribbling | Does it dribble after you have finished? | Y / N |
| 26. Double voiding | Do you pass urine, leave the toilet, and then have to go back again? | Y / N |
| **Pain symptoms** | | **Circle** |
| 27. Suprapubic pain | Do you get bladder pain felt over the pubic area? | Y / N |
| 28. Filling bladder pain | Does the b ladder gain worse as the bladder fills? | Y / N |
| 29. Voiding bladder pain | Do you get bladder pain during voiding? | Y / N |
| 30. Post void bladder pain | Do you get bladder pain after voiding? | Y / N |
| 31. Pain relieved by voiding | Do you get bladder pain fully relieved by voiding? | Y / N |
| 32. Partially voided relief | Do you get bladder pain partially relieved by voiding? | Y / N |
| 33. No voiding relief | Do you get bladder pain partially unrelieved by voiding? | Y / N |
| 34. Loin pain | Do you get flank pain in the kidney area? | Y / N |
| 35. Iliac fossa pain | Do you get pain to the right or left of the low abdomen? | Y / N |
| 36. Pain radiation to genitals | Do you get pain radiating into the vagina? | Y / N |
| 37. Pain radiation to legs | Do you get pain radiating down your legs? | Y / N |
| 38. Dysuria | Does it burn when you pass urine? | Y / N |
| 39. Urethral pain | Do you experience pain the urethra (the tube through which the urine passes? | Y / N |
